# Knowledge, Attitude, and Practice towards Antibiotic Use among the Support Staff of a Tertiary Care Teaching Hospital in India

**DOI:** 10.1101/2023.09.19.23295698

**Authors:** Nayana Nair, Neha Kadhe, Vrushali Badhan

## Abstract

**Aims & Objectives:** Antimicrobial resistance is a global problem arising mainly due to the irrational use of antibiotics, self-medication being one of the key contributors. Such practices are particularly common in developing countries where a large section of the population lacks awareness of the proper use of antibiotics.

Many studies in the past have assessed knowledge and practices about antibiotic use in public and healthcare personnel. However, literature on the Indian population, specifically, about support staff hospital workers (aya, hamal, ward boy, sweeper) is scarce. The fact that these workers closely interact with both doctors and patients renders them crucial to spreading the right information in the community. This study aims to assess the knowledge, attitude, and practices towards antibiotic use in the support staff of a tertiary healthcare hospital and to identify the demographic factors that affect their knowledge, attitude, and practices towards antibiotic use.

**Methodology:** A descriptive cross-sectional study was carried out using a self-administered, pretested, pre-validated structured questionnaire, in 403 support healthcare workers at a tertiary healthcare hospital. The questionnaire had 4 sections on demographic characteristics, knowledge of antibiotic use and resistance, attitude, and practices of antibiotic consumption.

Individual responses were scored and classified as good, average, or poor.

**Results:** Category-wise overall respondents’ scores were as follows: Knowledge (64.5% good, 27.8% average, 7.7% poor), Attitude (59.2% good, 33.9% average, 6.9% poor), and Practices (55.6% good, 44.1% average, 0.3% poor). Although 58% of respondents took antibiotics on doctor’s recommendation only, 93% did not use their leftover medication for family. 100% of people with age >50 years completed full courses of prescribed antibiotic therapy.

**Conclusion and Significance:** The study showed an association between good Knowledge, Attitude, and Practices with better education and income. While >50% study population scored well on all 3 criterion, certain issues, such as using leftover antibiotics for family and taking antibiotics without a doctor’s recommendation, are heavily prevalent. More awareness is required to prevent such practices.

## Introduction

The advent of antibiotics has been recognized as one of the greatest innovations that transformed medical treatment and outcome of infectious diseases. However, in recent decades faced the growing problem of emergence and spread of microbial resistance to antibiotics. ^[1-4]^ This presents a significant threat to public health globally by endangering their therapeutic effectiveness and increasing treatment failures leading to longer and more severe illness episodes with higher costs and mortality rates. Inappropriate and excessive use of antibiotics are among the key factors for increased spread of resistance. The improper use of antibiotics may arise from a complex interaction between numerous factors, such as prescribers’ knowledge and experiences, diagnostic uncertainty, perceptions of patients in relation to the patient-prescriber interaction, and insufficient patient education by physicians. ^[5-10]^

Irrational use of antibiotics also involves self-medication. The situation is serious in developing countries because of self-medication without prescription, over-the-counter sales of antibiotics, inadequate regulation of antibiotics, high cost of medical consultations, and dissatisfaction with medical practitioners. It is seen that more than 50% of antibiotics worldwide are bought without a prescription. ^[1-3]^ Over-the-counter purchase of antibiotics is a very common picture seen across our country where antibiotics are dispensed as a single dose therapy even for a milder illness like flu. The prevalence of self-medication with antibiotics in India reported in a study by *Muhammad Bilal et al* was 81.25% among the rural population of Sindh. ^[12]^

WHO reports of *September 2017* shows a serious deficiency of new antibiotics to combat antimicrobial resistance. The WHO Global Action on Antimicrobial Resistance Plan recognizes the role of societies in causing antibiotic resistance. Included in the list of possible causes of antibiotic resistance are the acts of inappropriate use of medicines that include failure to complete treatment, skipping of doses, re-use of leftover medicines, and overuse of antibiotics. The root cause is the lack of knowledge on the appropriate use of antibiotics. Several factors that are associated with public knowledge regarding antibiotic use have been reported to be demographic characteristics, including gender, age, education, family income, and place of residence. Moreover, social demographic characteristics and culture have been linked with practices towards antibiotics. Such information is important in designing future interventions needed to improve rational antibiotic use and ultimately restrain the dissemination of antibiotic resistance. ^[12-25]^

Many studies have been conducted to assess the knowledge, attitude, and practice about antibiotic use in public in different countries and have shown that the knowledge about the same is lacking among different sections of society. Studies have also been conducted to assess the same in the practicing doctors, healthcare personnel, and paramedical staff which includes resident doctors, interns, nurses, pharmacists, health officers, midwives, and laboratory technologists. ^[1-27]^ However, there is a scarcity of data about the knowledge, attitude, and practices of antibiotic use in the support staff hospital workers in India. This is the first study to our knowledge that has been done in this group.

In tertiary care hospitals, support staff workers such as aaya (female ward attendants), ward boy (male ward attendants), hamal (hospital porter), and sweeper (janitor), work in close association with clinicians. They have an opportunity to observe these clinicians and other healthcare workers and get influenced by their attitudes and practices. They can play an important role in spreading the right knowledge in the community. They may be approached in the community for health-related advice. Thus, it becomes imperative to test their knowledge, attitude, and practices. This study aims to assess the knowledge, attitudes, and practices about antibiotic use among such staff members of a tertiary care hospital and also to identify determinants of knowledge, attitudes, and practices regarding antibiotic use.

## Methodology

### Study Design, Population, and Sample size

A descriptive cross-sectional study was carried out using a self-administered, pretested, pre-validated structured questionnaire in 403 supporting healthcare workers - aya (female ward attendants), ward boy (male ward attendants), hamal (hospital porter), sweeper (janitor), at a tertiary healthcare hospital using a convenience sampling method after ethical approval from the institutional ethics committee. Written informed consent was taken from all participants. The sample size was calculated based on the population size of staff workers in tertiary care hospitals in Mumbai. ^[1-8]^

### Development of Questionnaire

The questionnaire was developed by reviewing available questionnaires in the literature (1-16). The questionnaire was designed in three language versions: English, Hindi, and Marathi. The questionnaire was originally developed in English, which was then translated into Marathi (the regional language of Maharashtra, India) and Hindi (the national language of India). Face and content validation of the questionnaire was done by two senior faculty members. Modifications were made based on the feedback provided. Then the questionnaire was pilot-tested among 20 respondents, who were not included in the final study analysis. Reliability showed internal consistency of the items tested with the Cronbachs α value of ≤ 0.8. ^[5]^

The questionnaire had 4 parts. ^[1-30]^ Part I recorded demographic characteristics - the respondents’ socio-demographic characteristics such as age, gender, educational level, employment, and residence. Part II was used to assess the knowledge of the participants about antibiotic use and resistance. Six questions were asked in this section, as shown in Table 3. One point was awarded for each correct response and zero for each wrong response, with a maximum obtainable correct score of six for each respondent. The knowledge score was divided into three levels indicated by poor (0-2), average (3-4), or good (5-6).

Part III contained eight questions (shown in Table 5) to assess the attitude. One point was awarded for each correct response and zero for each wrong response, with a maximum obtainable correct score of eight for each respondent. The attitude score was divided into three levels indicated by poor (less than 5), average (6-8), and good (more than 8).

Part IV gathered information regarding practices on antibiotic consumption (defined as antibiotic use within the past year) with the help of five questions (shown in Table 7). One point was awarded for each correct response and zero for each wrong response, with a maximum obtainable correct score of five for each respondent. The practice scores were divided into three levels indicated by poor (0-1), average (2-3), or good (4-5).

### Procedure

Support staff were screened. The questionnaire was administered to those eligible after taking written informed consent.

The inclusion criteria used were:

A. Adults aged 18 years and above,
B. Able to understand either of the three languages-English, Marathi, or Hindi and
C. Aware of the term antibiotics.

Each member was given the questionnaire in the language he/she is comfortable with (Marathi or Hindi). If the person was not able to read, questions were read out to them in person and the questionnaire was filled.

## Statistics

Descriptive statistics were calculated using frequencies and percentages for categorical variables. The significance of the impact of demographic parameters on the scores of KAP questions was determined by the p-value obtained from Chi-square and Fisher’s exact test.

We considered participants who provided a >90% filled questionnaire. The total sample size (N) may not tally for all demographic parameters and questions due to missing values for some respondents.

Table 1 lists the demographic of the respondents, categorized by Gender, Education, Age, and Monthly Income. All category labels are ranked from highest to lowest for the corresponding parameter, for example, IC1 indicates the highest income group and IC4 indicates the lowest income category.

**Table 1.** Demographic characteristics of respondents, N = 392.

| Parameters | Category Labels | Categories | Frequency (%) |
| --- | --- | --- | --- |
| **Gender** | M | Male | 194 (49.5%) |
|  | F | Female | 198 (50.5%) |
| **Age** | AC1 | >60 | 29 (7.4%) |
|  | AC2 | 50-59 | 142 (36.3%) |
|  | AC3 | 40-49 | 121 (30.9%) |
|  | AC4 | 30-39 | 88 (22.5%) |
|  | AC5 | 18-29 | 11 (2.9%) |
| **Education** | EC1 | Graduate and above | 168 (43.2%) |
|  | EC2 | Higher secondary passed | 87 (22.4%) |
|  | EC3 | Secondary passed | 38 (9.7%) |
|  | EC4 | Primary | 44 (11.3%) |
|  | EC5 | Uneducated | 52 (13.4%) |
| **Monthly income (INR)** | IC1 | >30000 | 89 (22.8%) |
|  | IC2 | 15000-29999 | 135 (34.6%) |
|  | IC3 | 10000-14999 | 89 (22.8%) |
|  | IC4 | <10000 | 77 (19.8%) |

Total sample size (N) may not tally for all parameters due to missing values for some respondents.

## Results

Of the 636 respondents screened, only 403 (63.36%) were aware of the term antibiotics and that they were used for the treatment of infections cough, or cold fever. Only 86.41 % of these, in addition, knew that antibiotics are used in the treatment of bacteria and not viruses.

A total of 392(61.6%) questionnaires were analyzed in this study; the remaining 11 incomplete questionnaires (<90% filled) were excluded. In terms of participants’ occupation, 198(50.5%) were ayas (female ward attendants), 93(23.7%) ward boys (male ward attendants), 56(14.3%) sweepers (janitor), and 45(12.2%) were hamals (hospital porter). Demographic characteristics of respondents are given in Table 1.

Table 2 shows the overall KAP scores of the respondents. We observed that 250(64.5%), 232(59.2%), and 218(55.6%) respondents scored “good” in the Knowledge, Attitude, and Practices sections, respectively. However, the percentage of correct responses recorded for individual questions exhibited large variability (tables 3, 5, and 7). 155(39.7%) respondents had incorrect knowledge about antibiotics having minimal side effects (KQ3, table 3).

**Table 2.** Number (and percentage) of Respondents with Good, Average and Poor KAP Scores, N = 392.

| Grade | Knowledge (%) | Attitude (%) | Practice (%) |
| --- | --- | --- | --- |
| **Good** | 250 (64.4) | 232 (59.2) | 218 (55.6) |
| **Average** | 108 (27.9) | 133 (33.9) | 173 (44.1) |
| **Poor** | 30 (7.7) | 27 (6.9) | 1 (0.3) |

Total sample size (N) may not tally due to missing values for some respondents.

**Table 3.** Frequency of Correct Responses to individual questions for Knowledge assessment.

| Question IDs | Knowledge Questions | Correct responses | Incorrect responses |
| --- | --- | --- | --- |
| KQ1 | Antibiotics are used in the treatment of infections with bacteria and not with viruses. | 86.4% | 13.6% |
| KQ2 | Antibiotics can destroy the organisms which are good for our body | 85.9% | 14.1% |
| KQ3 | Antibiotics are safe drugs with minimal side effects. | 60.3% | 39.7% |
| KQ4 | Different antibiotics are needed to cure different types of diseases. | 80.8% | 19.2% |
| KQ5 | Antibiotics that are able to kill an organism, may not be able to do so later if organism develops resistance. | 80.8% | 19.2% |
| KQ6 | Improper use of antibiotics (improper dose and duration of therapy) can lead to drug resistance. | 83.9% | 16.1% |

Some attitude assessment questions also garnered a high percentage of incorrect responses (table 3). Specifically, many respondents thought that antibiotics were to be consumed for any sickness, and were okay with procuring them from a chemist without a prescription and stocking them for future needs (AQ1, AQ3, and AQ5). A casual attitude towards adhering to the prescribed regimen was also observed as 99(25.4%) and 90(23.1%) respondents found it okay to stop the antibiotics upon feeling better and to tinker with the prescribed dose, respectively.

The practices followed by the respondents as assessed by the scored questions, are depicted in Table 7. Some of the practices observed were interesting. 224(57.7%) respondents took antibiotics on a doctor’s recommendation; but, 48(12.4%) respondents took self-medications, 40(10.3%) took them with a non-doctor, healthcare worker’s advice (nurse, etc.), 59(15.2%) took medications on friends/family member’s advice and 17(4.4%) on chemist’s recommendation.

Apart from what is illustrated in Table 7, we also observed that 364(92.9%) respondents had consumed antibiotics at least once in the last 1 year; 202(51.5%) respondents have used antibiotics 1-2 times, 128(32.7%) respondents have used antibiotics 3-4 times and 34(8.7%) had used them 5-6 times. 27(6.9%) had not taken antibiotics in the last 1 year. 193(49.4%) respondents had taken antibiotics as recently as the last 3 months, 119(30.5%) respondents took antibiotics 3-6 months ago, 35(8.9%) respondents had taken antibiotics between 6 months and 1 year and 42(10.8%) had taken them more than a year ago.

Antibiotics were commonly used for the following conditions: 35(9%) flu, 175(44.8%) diarrhoea, 237(60.6%) cough, 217(55.5%) ear infection, 142(36.3%) sore throat, 49(12.5) common cold, 2(0.5%) acidity, 1(0.3%) bodyache, 1(0.25%) tooth pain. 267(68.2%) respondents took 3-5 days course of antibiotics, 86(22%) took for less than 3 days, 38(9.7%) took them for 7-10 days.

Tables 4, 6, and 8 show the variation in knowledge, attitude, and practice across different demographic categories. For brevity and clarity of presentation, some questions in the attitude section have been omitted from Table 6. As expected, higher education and income groups typically recorded a relatively larger frequency of correct responses to most of the KAP questions. However, table 6 shows that more educated and high-income respondents had a worse attitude towards stocking antibiotics for themselves or family members. We also observed that respondents >50 years old were more likely to complete the recommended therapy.

**Table 4.**
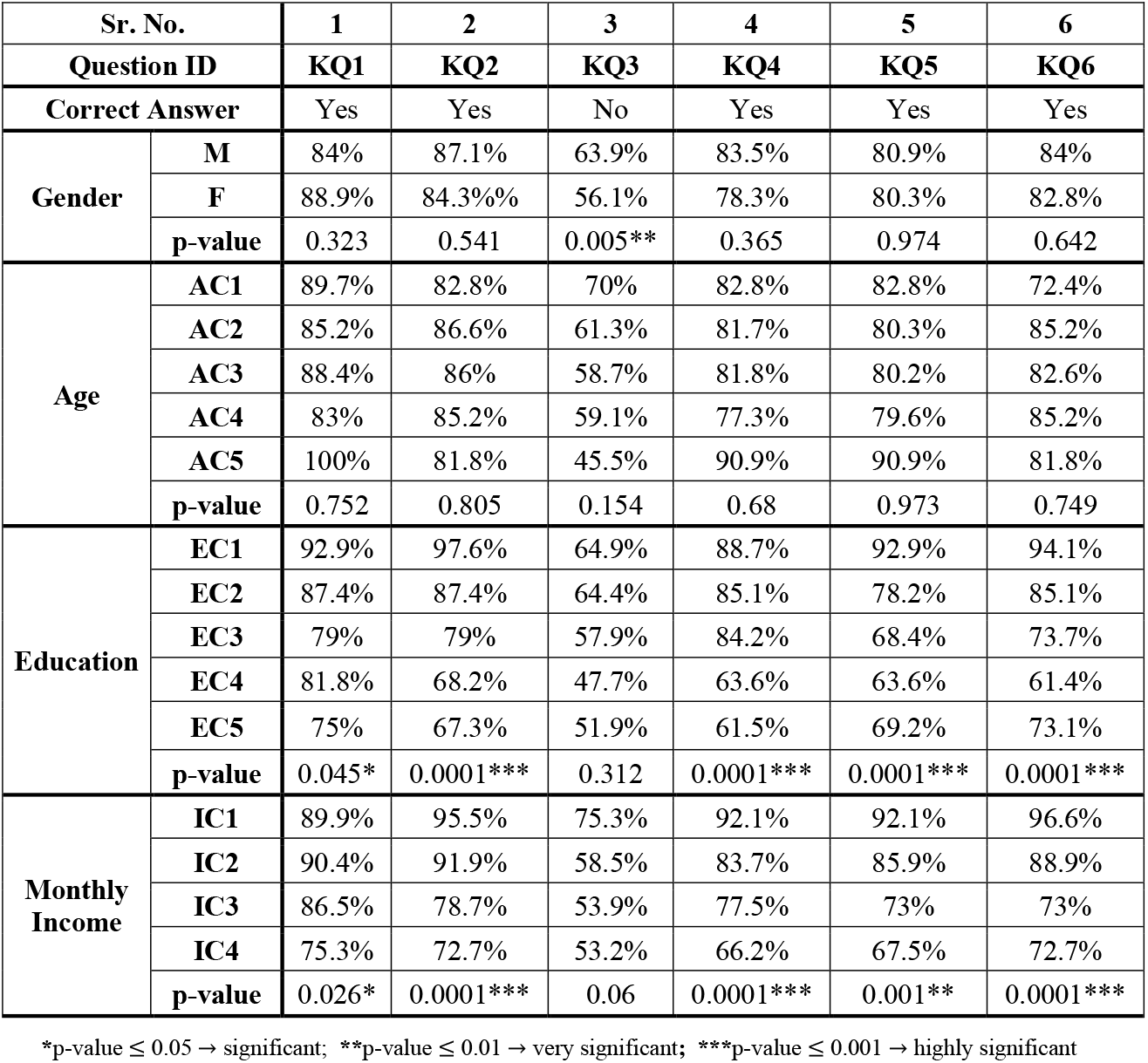
Percentage of correct responses to Knowledge assessment questions in various Demographic categories.

| Sr. No. |  | 1 | 2 | 3 | 4 | 5 | 6 |
| --- | --- | --- | --- | --- | --- | --- | --- |
| Question ID |  | KQ1 | KQ2 | KQ3 | KQ4 | KQ5 | KQ6 |
| Correct Answer |  | Yes | Yes | No | Yes | Yes | Yes |
| Gender | M | 84% | 87.1% | 63.9% | 83.5% | 80.9% | 84% |
|  | F | 88.9% | 84.3% | 56.1% | 78.3% | 80.3% | 82.8% |
|  | p-value | 0.323 | 0.541 | 0.005** | 0.365 | 0.974 | 0.642 |
| Age | AC1 | 89.7% | 82.8% | 70% | 82.8% | 82.8% | 72.4% |
|  | AC2 | 85.2% | 86.6% | 61.3% | 81.7% | 80.3% | 85.2% |
|  | AC3 | 88.4% | 86% | 58.7% | 81.8% | 80.2% | 82.6% |
|  | AC4 | 83% | 85.2% | 59.1% | 77.3% | 79.6% | 85.2% |
|  | AC5 | 100% | 81.8% | 45.5% | 90.9% | 90.9% | 81.8% |
|  | p-value | 0.752 | 0.805 | 0.154 | 0.68 | 0.973 | 0.749 |
| Education | EC1 | 92.9% | 97.6% | 64.9% | 88.7% | 92.9% | 94.1% |
|  | EC2 | 87.4% | 87.4% | 64.4% | 85.1% | 78.2% | 85.1% |
|  | EC3 | 79% | 79% | 57.9% | 84.2% | 68.4% | 73.7% |
|  | EC4 | 81.8% | 68.2% | 47.7% | 63.6% | 63.6% | 61.4% |
|  | EC5 | 75% | 67.3% | 51.9% | 61.5% | 69.2% | 73.1% |
|  | p-value | 0.045* | 0.0001*** | 0.312 | 0.0001*** | 0.0001*** | 0.0001*** |
| Monthly Income | IC1 | 89.9% | 95.5% | 75.3% | 92.1% | 92.1% | 96.6% |
|  | IC2 | 90.4% | 91.9% | 58.5% | 83.7% | 85.9% | 88.9% |
|  | IC3 | 86.5% | 78.7% | 53.9% | 77.5% | 73% | 73% |
|  | IC4 | 75.3% | 72.7% | 53.2% | 66.2% | 67.5% | 72.7% |
|  | p-value | 0.026* | 0.0001*** | 0.06 | 0.0001*** | 0.001** | 0.0001*** |

\*p-value $\leq 0.05$ → significant; \*\*p-value $\leq 0.01$ → very significant; \*\*\*p-value $\leq 0.001$ → highly significant

**Table 5.**
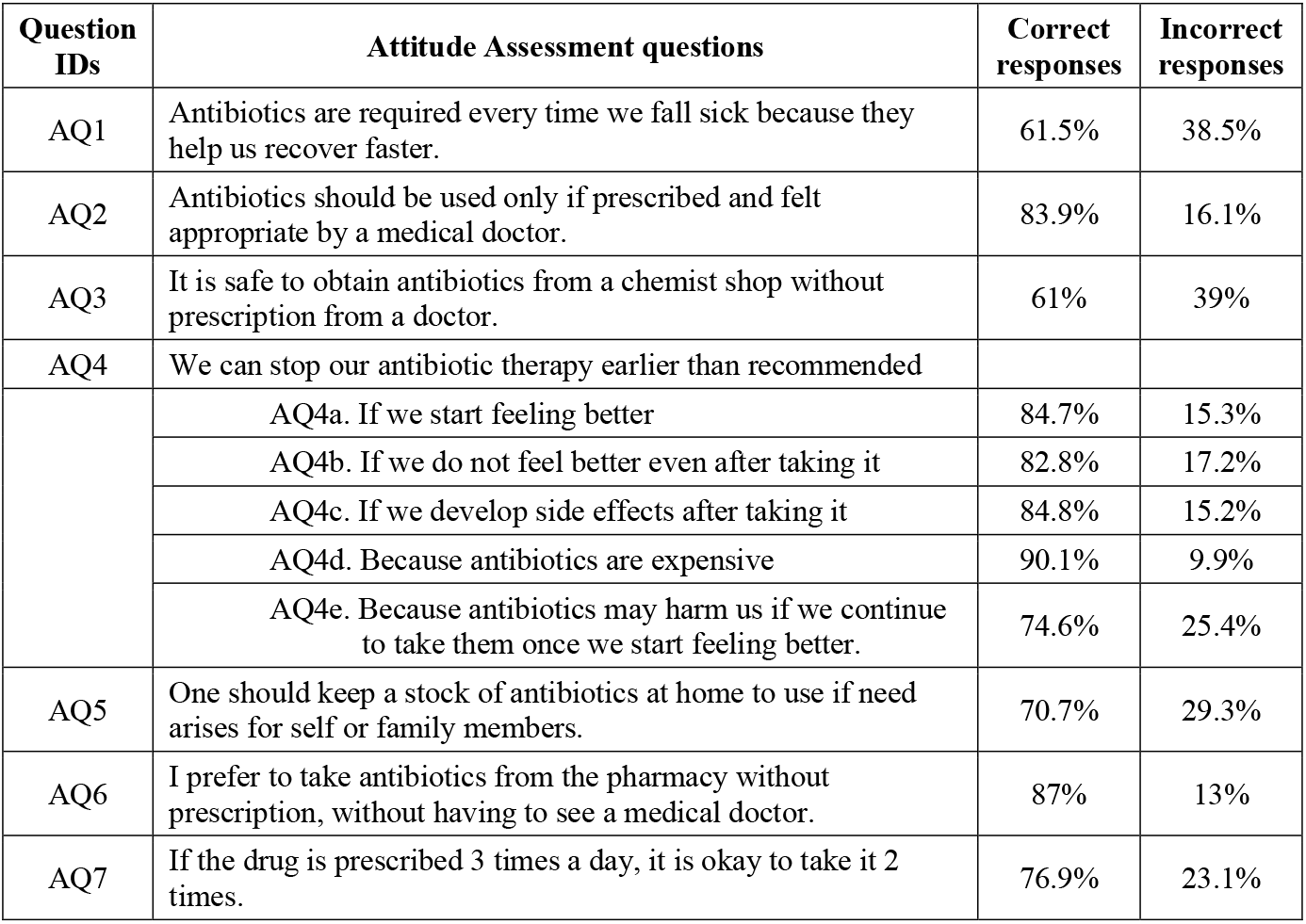
Frequency of Correct Responses to individual questions for Attitude assessment.

| **Question IDs** | **Attitude Assessment questions** | **Correct responses** | **Incorrect responses** |
| --- | --- | --- | --- |
| AQ1 | Antibiotics are required every time we fall sick because they help us recover faster. | 61.5% | 38.5% |
| AQ2 | Antibiotics should be used only if prescribed and felt appropriate by a medical doctor. | 83.9% | 16.1% |
| AQ3 | It is safe to obtain antibiotics from a chemist shop without prescription from a doctor. | 61% | 39% |
| AQ4 | We can stop our antibiotic therapy earlier than recommended |  |  |
|  | AQ4a. If we start feeling better | 84.7% | 15.3% |
|  | AQ4b. If we do not feel better even after taking it | 82.8% | 17.2% |
|  | AQ4c. If we develop side effects after taking it | 84.8% | 15.2% |
|  | AQ4d. Because antibiotics are expensive | 90.1% | 9.9% |
|  | AQ4e. Because antibiotics may harm us if we continue to take them once we start feeling better. | 74.6% | 25.4% |
| AQ5 | One should keep a stock of antibiotics at home to use if need arises for self or family members. | 70.7% | 29.3% |
| AQ6 | I prefer to take antibiotics from the pharmacy without prescription, without having to see a medical doctor. | 87% | 13% |
| AQ7 | If the drug is prescribed 3 times a day, it is okay to take it 2 times. | 76.9% | 23.1% |

**Table 6.** Percentage of correct responses to Attitude assessment questions in various Demographic categories.

| Sr. No. |  | 1 | 2 | 3 | 4 |  | 5 | 6 | 7 |
| --- | --- | --- | --- | --- | --- | --- | --- | --- | --- |
| Question ID |  | AQ1 | AQ2 | AQ3 | AQ4c | AQ4e | AQ5 | AQ6 | AQ7 |
| Correct Answer |  | No | Yes | No | No | No | No | No | No |
| Gender | M | 65.5% | 85.1% | 66.5% | 86.6% | 75.3% | 74.1% | 88.1% | 74.6% |
|  | F | 57.1% | 82.7% | 55.6% | 82.7% | 73.6% | 67.2% | 85.9% | 79.3% |
|  | p-value | 0.226 | 0.804 | 0.077 | 0.492 | 0.918 | 0.08 | 0.484 | 0.317 |
| Age | AC1 | 58.6% | 82.8% | 69% | 96.6% | 96.5% | 86.2% | 86.2% | 82.8% |
|  | AC2 | 66.9% | 86.5% | 63.4% | 88% | 70.4% | 64.5% | 87.3% | 73.8% |
|  | AC3 | 55.4% | 81.8% | 60.3% | 83.5% | 75.8% | 71.9% | 91.7% | 76% |
|  | AC4 | 60.2% | 83% | 55.7% | 77% | 72.7% | 71.6% | 79.6% | 80.7% |
|  | AC5 | 63.6% | 81.8% | 54.6% | 90.9% | 72.7% | 81.8% | 90.9% | 81.8% |
|  | p-value | 0.744 | 0.586 | 0.833 | 0.019* | 0.569 | 0.0001*** | 0.483 | 0.536 |
| Education | EC1 | 67.3% | 88% | 67.9% | 86.9% | 75% | 67.3% | 88.7% | 81% |
|  | EC2 | 69% | 80.5% | 60.9% | 81.4 | 81.6% | 69% | 83.9% | 75.6% |
|  | EC3 | 52.6% | 86.8% | 76.3% | 73.7% | 70.3% | 73.7% | 92.1% | 76.3% |
|  | EC4 | 54.6% | 81.8% | 43.1% | 81.8% | 68.2% | 75% | 77.3%% | 72.7% |
|  | EC5 | 42.3% | 75% | 46.2% | 92.3% | 71.2% | 78.4% | 90.4% | 69.2% |
|  | p-value | 0.004** | 0.312 | 0.006** | 0.215 | 0.076 | 0.783 | 0.246 | 0.051 |
| Monthly Income | IC1 | 75.3% | 95.5% | 79.8% | 85.4% | 75.3% | 67.4% | 88.8% | 83.2% |
|  | IC2 | 64.4% | 91.9% | 59.3% | 81.3% | 74.1% | 64.4% | 83.7% | 77% |
|  | IC4 | 48.1% | 72.7% | 50.7% | 89.6% | 75.35 | 77.6% | 89.6%% | 74% |
|  | IC5 | 55.1% | 78.7% | 53.9% | 85.4% | 73.9% | 77.5% | 87.6% | 72.7% |
|  | P value | 0.0001*** | 0.0001*** | 0.002** | 0.342 | 0.614 | 0.043* | 0.697 | 0.246 |

\*p-value $\leq 0.05$ → significant; \*\*p-value $\leq 0.01$ → very significant; \*\*\*p-value $\leq 0.001$ → highly significant

**Table 7.** Frequency of Correct Responses to individual questions for Practice assessment.

| Question IDs | Practice Assessment Questions | Correct responses | Incorrect responses |
| --- | --- | --- | --- |
| PQ1 | Antibiotic taken on doctor's recommendation | 57.7% | 42.3% |
| PQ2 | Have you taken an antibiotic in the last one year even if your doctor told you that there is no need for you to take it? | 96.9% | 3.1% |
| PQ3 | Have you stopped an antibiotic therapy earlier because you found the treatment expensive? | 98.5% | 1.5% |
| PQ4 | I have always completed the full recommended duration of days of therapy of antibiotics. | 97.9% | 2.1% |
| PQ5 | I have used my left-over antibiotics for my family member if they fall sick. | 6.9% | 93.1% |

**Table 8.**
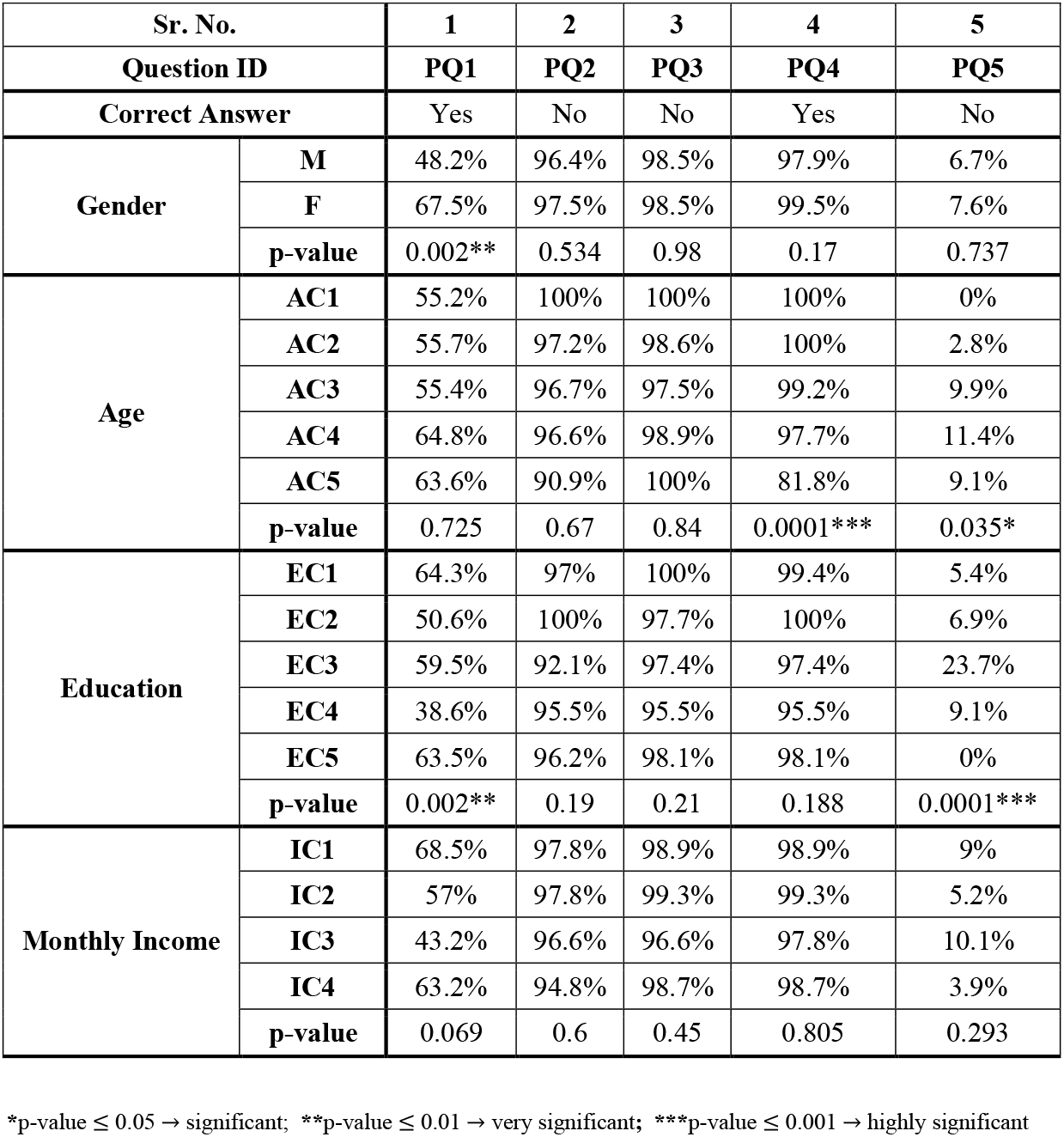
Percentage of correct responses to Practice assessment questions in various Demographic categories.

## Discussion

This study aims to assess the knowledge, attitude, and practice regarding antibiotic use in healthcare support staff like aaya (female ward attendants), ward boy (male ward attendants), hamal (hospital porter), and sweeper (janitor), as they work in close association with clinicians. Previous studies on KAP have looked at doctors, nurses, and medical students. ^[9-30]^ These people have varying degrees of formal training for antibiotic use, but support staff do not have formal training. However, they are in a position to influence KAPs of the community as they come in close association with them. Hence, it’s important to assess their KAP.

Some studies in other countries such as Malaysia have evaluated the kap of support staff however to the best of our knowledge no such studies have been done in the Indian population. ^[1]^

The screening questions revealed that 36.6% of the screened staff were unaware of the term antibiotics and their role. Of the 63.4% who correctly answered the screening questions (i.e. they were aware of the term antibiotics and felt they were used in the treatment of infections/cold, cough, fever), 64.4%, 59.2%, and 55.6% showed good knowledge, attitude, and practice respectively. Less education and low income were associated with poor scores in knowledge about antibiotics. Only 86.4% of those felt that antibiotics were used for the treatment of infections or cough, cold, and fever, in addition, knew that antibiotics are used for the treatment of bacterial infections and not viral ones. Further, 21.5% had used antibiotics for the common cold and flu in the last 1 year. The lack of knowledge about the use of antibiotics in bacterial or viral infections amongst the respondents might have resulted in the use of antibiotics for viral fever, most probably via self-medication or on recommendation by a paramedical healthcare worker or a friend/family member. This further highlights the importance of educating paramedical healthcare workers about antibiotic use. ^[1-3]^

38.5% believed that antibiotics are required every time we fall sick because they help us recover faster. Lower education and low income were associated with this belief. Further, 39.7% of the respondents said that antibiotics are safe drugs with minimal side effects, but 85.9% were aware that antibiotics can destroy organisms good for the body; 83.9% were aware that improper use of antibiotics can lead to drug resistance; and 80.8% also knew that a sensitive organism can become resistant to the antibiotic at a later date. This knowledge might be due to their close association with doctors in the work setting. ^[1]^

A study conducted in Malaysia showed 45.6% of respondents opted to stop an antibiotic course midway if symptoms improved. ^[1]^ In an Indian study analyzing doctors, interns, and nurses, only 7.1% of respondents answered affirmatively to stopping the antibiotic course midway when symptoms improve. ^[6, 18, 29]^ In our study, 15.3% of respondents said it is okay to stop taking antibiotics if we start feeling better. However, in practice, 98.7% of respondents reported having completed the full recommended course of antibiotic therapy. This might be because when people fall sick, they are more concerned and show better adherence.

83.9% felt that antibiotics should be used only when prescribed and recommended by the doctor. However, 39% believed it was safe to obtain antibiotics from the chemist shop without prescription from a doctor. In practice, only 57.7% had taken the antibiotics on the recommendation of the doctor. The remaining had taken them as self-medication (12%), or on the recommendation of a paramedical healthcare worker (10%), friends/family member (15%), or chemist (4.4%). Despite the new law prohibiting the dispensing of antibiotics by chemists without a valid prescription, 4.4 % of respondents had taken the antibiotics on the recommendation of the chemist. It becomes imperative to highlight through training that antibiotics should be used only if deemed necessary by the physician. More females took antibiotics on the doctor’s recommendation. This could point towards a lower tendency to self-medication amongst females. ^[6]^

93.1% of the respondents had consumed antibiotics in the last 1 year; of which 51.5% had taken them 1-2 times and 41.3% had taken them 3-6 times in the last year. 6.9% had not taken antibiotics in the last 1 year. In a separate question, 10.8% said they took antibiotics more than a year ago. This discrepancy might be due to recall bias. 41.3% of respondents taking antibiotics 3-6 times a year highlights the concern of overuse. ^[1]^

44.8% of respondents had taken antibiotics for diarrhea. It would have been interesting to find out if these were indeed of bacterial origin and whether given as an irrational FDC. 22% of respondents had taken the antibiotics for just 1-2 days but 98.7% said that they have always completed the full recommended duration of days of antibiotic therapy. This points to irrational prescribing by physicians or wrong advice by non-doctors. This also highlights the influence of other people, namely the paramedical healthcare workers, lay people, and the chemist, on deciding the duration of therapy, especially if the antibiotics are taken on their recommendation. Though 70.7% of respondents felt we should not keep a stock of antibiotics at home if the need arises for use by self or a family member, 93.1% of respondents had used their left-over antibiotics for their family members when they fell sick. Thus, there is a discrepancy between attitude and actual practice. It would be interesting to know if this was through self-medication or otherwise. ^[1-3]^

Overuse of antibiotics has been observed in the survey; 38.5% think we should take antibiotics every time we fall sick, and 21.5% use them for the common cold and flu. 41.3% had taken them 3-6 times in the last year. This points to a Knowledge gap but also could be irrational prescribing. A change in attitude is also required as 23.1% of respondents felt that it is ok to skip an antibiotic dose.

Respondents with high income and higher education had better knowledge and attitude scores. However, practices assessed by questions PQ1 and PQ5 of Table 8, regarding self-medication and using leftover antibiotics for family members, respectively, were very poor in all categories. This highlights the need to spread awareness about good antibiotic use practices in the community.

## Conclusion

It was concluded that knowledge, attitude, and practice regarding antibiotic use among support staff hospital workers in India was good. However, it is alarming that 42% still had taken antibiotics in the last year without a doctor’s prescription.

Though respondents had good knowledge scores, there is a gap in the knowledge in certain areas which could be the source of discrepancy between attitudes and actual practices. We should develop a good culture of antibiotic practices at individual, institutional, and regulatory levels.

## Data Availability

All data produced in the present work are contained in the manuscript.

## Notes

### Competing Interest Statement

The authors have declared no competing interest.

### Funding Statement

This study did not receive any funding.

### Author Declarations

Ethics committee of Lokmanya Tilak Municipal General Hospital and Medical College, Mumbai, India Study Reference ID: 2016-04146

